# Remote monitoring of depression severity: A machine learning approach

**DOI:** 10.1101/2023.08.22.23294431

**Authors:** Vincent Holstein, Habiballah Rahimi-Eichi, Daniel Emden, Lara Gutfleisch, Alexander Refisch, Janik Goltermann, Ramona Leenings, Nils Winter, Tilo Kircher, Igor Nenadić, Ronny Redlich, Elisabeth Johanna Leehr, Katharina Dohm, Justin Baker, Udo Dannlowski, Nils Opel, Tim Hahn

## Abstract

Depression is a widely prevalent psychiatric illness with variable levels of severity that necessitate different approaches to treatment. To enhance the management of this condition, there is a growing interest in utilizing mobile devices, especially smartphones, for remote monitoring of patients. This study aims to build prediction models for depression severity based on active and passive features collected from patients with major depressive disorder (MDD) and healthy controls to assess the feasibility of remote monitoring of depression severity.

Using data from 142 participants (85 healthy controls, 67 MDD) we extracted features such as GPS-derived mobility markers, ecological momentary assessments (EMA), age, and sex to develop machine learning models of depression severity on the different diagnostic subgroups in this cohort.

Our results indicate that the employed models outperformed baseline estimators in random split scenarios. However, the improvement was marginal in user-split scenarios, highlighting the need for larger and more diverse samples for clinical utility. Among the features, mood EMA emerged as the most influential predictor, followed by GPS-derived mobility features. Models also showed a significant association between depression severity and average reported mood, as well as GPS-derived mobility markers such as number of places visited and percent home.

While predicting composite depression scores is important, future studies could explore predicting individual symptom items or symptom groups for a more comprehensive assessment of depression severity. Challenges for clinical utility include participant dropout, which could be addressed through more engaging app design to promote user adherence. Harmonization of phone-derived measures is also crucial to facilitate model transfer across studies.

In conclusion, this study contributes valuable evidence supporting the potential utility of smartphone data for mood state monitoring and predicting depression severity. Future research should focus on predicting depression further ahead in time and addressing the challenges identified to create more robust and effective depression monitoring solutions using smartphone-based data.

## Introduction

Depression is a widespread psychiatric illness that affects millions of individuals worldwide, with a lifetime prevalence of 20% [1], projected to be the lead contributor to global disease burden by the year 2030 [2]. Being an ever-increasing burden on our healthcare systems, there is an increasing need for improved monitoring and predictive tools to enhance the management of this condition. As relying solely on human judgement is costly and subject to potential biases attention in recent years has turned to mobile devices for the quantitative assessment of behaviour [3, 4]. Sometimes referred to as digital phenotyping smartphones and mobile technology have opened up new possibilities for remote monitoring and data collection, providing valuable insights into the daily lives of individuals with depression [5, 6].

Since the inception of the iPhone in 2008 the prevalence of smartphones has rapidly increased with smartphone penetration reaching >80% in the United States and 50% worldwide [7, 8]. The widespread use of smartphones has made them a prime target for digital phenotyping efforts, with multiple uses in a range of different mental health conditions such as Bipolar disorder, OCD and MDD [9–11]. Smartphones are often used to collect both active measures, such as momentary ecological assessments and passive measures, such as GPS-derived mobility markers, device usage or app usage. A key challenge for the application of remote monitoring via smartphones is the reliable prediction of depression severity for in-time intervention and improved treatment monitoring. In previous studies of depressed populations, multiple different smartphone-derived measurements have been used such as social interactions, location, activity measures, EMAs and phone use [11]. For EMAs, it has been shown that depressed individuals report lower average mood ratings [12, 13]. These EMA-reported mood ratings are also significantly associated with composite scores of depression such as PHQ-9 [14, 15]. Similarly, GPS-derived mobility markers such as percent home or the number of locations visited have been shown to be altered in depression [16], while also correlating strongly with depression severity over time [17, 18]. Daily GPS features and mood do not seem to be strongly related however, with substantial day-to-day variability [16, 17].

These promising findings have prompted researchers to harness smartphone-derived sensor data for the prediction of psychiatric diagnoses, utilizing an array of sensor-based features. Multiple studies have effectively classified individuals based on their depression status, achieving accuracies ranging from 0.60 to over 0.85 [19–22]. These studies utilized a range of different features such as GPS, screen status or internet connectivity. Beyond diagnosis prediction, some investigations have attempted to forecast depression severity through smartphone data. While certain studies, such as those by Asare et al. and Ware et al., focused on predicting individual symptoms using classifiers, others, including Saeb et al., Wang et al., Pedrelli et al., and Lewis et al. have predicted depression severity on a continuous scale [17, 23, 24]. For continuous predictions, the PHQ-9 and Hamilton Depression Rating Scale (HAM-D) were commonly used with a subset of studies incorporating alternative metrics like the PHQ-8. Notably, varying model performances were reported, with some studies yielding inconclusive outcomes at the population level [16]. It is noteworthy that the majority of these models relied on passive sensing measures, with only a limited number incorporating active measures such as Ecological Momentary Assessments (EMAs).

Contributing to a growing body of evidence, our study constructs prediction models for depression severity by leveraging both active and passive features obtained from individuals with major depressive disorder (MDD) and healthy controls. Through the utilization of permutation feature importances and statistical modelling techniques, we not only replicate insights from prior research but also introduce supplementary evidence by addressing the integration of missing data within our machine learning frameworks. We are hopeful that the outcomes of our research will help to guide forthcoming investigations in the realm of digital phenotyping.

## Methods

The primary goal of the study was to analyze the feasibility of mood prediction from active and passive smartphone data in a clinical population. We also analyzed the statistical relationships of the different predictors to our target to inform potential future studies. In this section we first describe our cohort and then our preprocessing and analysis methods used in this study.

### Subjects

N=1550 Participants from the ReMAP study with available GPS data, demographic information, ecological momentary assessments and sleep questionnaires from the Remote Monitoring in Psychiatry (ReMAP) study with data between January 2019 and September 2022 were selected for further processing. All study participants were recruited as part of ongoing longitudinal cohort studies. For more information on these studies please refer to the supplementary material. After pre-processing quality control, processing and post-processing quality control 207 Participants remained, of which 156 were either healthy controls (N=96) or diagnosed with major depressive disorder (N=60). Patients were recruited and assessed at the Institute for Translational Psychiatry, Department of Psychiatry at the University of Muenster. Current and lifetime psychopathological diagnoses were assessed using the Structured Clinical Interview for DSM-IV (SCID-I). Patients included those with mild, moderate, severe and (partially) remitted MDD. All subjects were educated about study aims, data collection, data security and financial compensation. The study was approved by the local institutional review board and Informed consent was obtained from each subject. Further information about study design can be found in the supplement as well as our previous work [25, 26].

### Quality Control

Geolocation data underwent quality control before (pre-processing quality control) and after processing (post-processing quality control). For pre-processing quality control, a custom script was used that automatically discarded participants with less than 30 days of data or with a minimum timestamp difference >6 hours, indicating low data density. We also discarded all data points during periods where individual participants were isolated at home due to COVID-19 infections. After this automated quality control step accuracy data over time was plotted and visually inspected. If there were missing periods for >30% of collection time or other anomalies data was discarded. After processing daily location maps from the DPLocate pipeline were plotted and visually inspected. If anomalies were detected data was discarded and did not enter the final statistical analysis.

### Geolocation Data Processing

Geolocation was processed using the DPLocate pipeline. Data were converted to Matlab format, signal epochs were extracted using a threshold of 1 datapoint and temporally filtered. Points of interest (POIs) were extracted using the modified Density-Based Clustering Algorithm with Noise (DBSCAN), calculating the 80 most visited locations with the maximum number of epochs in their 50 meters neighbourhood and then pruning overlapping clusters to find the most visited locations [27, 28]. Epochs were assigned to POIs, the home location was estimated as the most commonly visited location and metrics of interest were calculated from these epochs. These metrics were: Average Distance from home, Radius of Mobility, Percent Home and the number of places visited.

The pipeline was run using MatLab 2019a.

### Psychometric Questionnaire Data

For the assessment of depressive symptoms, the German version of the Beck Depression Inventory (BDI) was used [29]. The questionnaire was administered every 2 weeks through the ReMAP phone app as specified by the study protocol. For temporal granularity, a daily ecological momentary assessment of mood and sleep consisting of two items with a sliding scale (0-10) was delivered through the study app at random times between 8 am and 10 pm. Users were notified via push notifications. Both the BDI and EMA questionnaires have been externally validated in our previous work [25, 26].

### Predictive Data Processing

For our predictive models, the data was averaged over the 2-week period prior to BDI administration. 2-week windows with less than 5 data points were labelled as missing and assigned a missing value of 1 for that time window. This was done to model missingness within the data while preserving an adequate sample size. Predictors were GPS-derived mobility markers (percent home, number of places visited, radius of mobility, distance from home), ecological momentary assessments (mood, sleep quality), age, sex and phone platform.

### Predictive Modelling

All machine learning models were developed using the PHOTONAI package [30]. Four different predictive models were used: Random Forest, Automatic Relevance Determination Regression, Huber Regression and a Support Vector Machine. Every model was evaluated using a nested 10×10-Fold cross-validation with the best model being selected on mean absolute error and grid search as an optimizer strategy. Additionally, mean squared error, Spearman rank correlation and Pearson correlation were calculated. We also calculated a baseline estimate (referred to as a “Dummy Estimator”) by predicting the sample mean. After pipeline training and evaluation we calculated the permutation feature importance to assess the importance of different predictors. To evaluate our predictive models under different conditions we trained each pipeline on two different data splits. First, we split the data randomly (random split), leading to data from most participants being available in the training and test set. This was done to model a scenario where some user data is already available for model tuning. Second, we also performed a user split where some users were held out in the test set and not seen during training. This was done to estimate how well the model might generalize to new unseen participants. A potential time split was not performed due to the high variability in the amount of data points per user.

### Inferential Modelling

To gain insight into the nature of the statistical relationships within the data we built linear regression models for the different predictors. We modelled the relationship between BDI and different predictors using linear mixed models with a random intercept for each subject with age and sex as covariates. Modelling the group relationship we used the averaged values of the predictors in a linear regression. All statistical analyses were performed using the statsmodels Python package.

## Results

### Prediction Model

To predict the BDI sum score from mobile data we we trained each estimator model on either patients with major depressive disorder (MDD), healthy controls (HC) only or both of them combined (Comb). 142 subjects went into the analysis with 85 HC and 57 MDD patients. The age range spanned 18 to 67 years with a median age of 35 years. Demographic information on each subgroup can be found in the supplementary materials. BDI was predicted in each of the subgroups using both a random split and user split, leading to 24 different classifier pipelines, excluding the Dummy estimator.

Predicting BDI score in the MDD subgroup was best achieved by the Random Forest in the random split cohort with a mean absolute error of 5.301 and a mean squared error of 44.543. However, the other classifiers showed a highly similar predictive performance, with no classifier being clearly superior. Each classifier performed better than the dummy estimator with a mean absolute error of 7.25 In the user split the classifiers performed even more similarly with no clear best classifier. Here the estimators performed only slightly better than the dummy estimator (Mean absolute error: 7.59). For each pipeline, we also performed a permutation test to quantify whether the correlation between the true and predicted values was due to chance. All models performed significantly above chance with p-values <0.001. Predicting BDI score in the HC subgroup was again best achieved by the Random Forest in the random split subgroup with a mean absolute error of 3.793 and a mean squared error of 27.101. All estimators performed better than the dummy estimator (Mean absolute error: 5.962). In the user split subgroup, the results between the estimators were much more similar with the SVR ranking as the best classifier, however, most classifiers still performed better than the dummy estimator (Mean absolute error: 6.139). The permutation test was again significant for each model (p<0.001).

Predicting BDI score in the combined group was again best achieved by the random forest when using a random split with a mean absolute error of 3.793 and a mean squared error of 27.101. All estimators outperformed the Dummy (Mean absolute error: 5.962). The SVR was the best estimator in the group split with a mean absolute error of 4.259 and a mean squared error of 32.921. In the combined group all estimators performed highly similarly both in the random and the user split condition. In the combined condition all models performed above chance (p<0.001)

### Feature importances

For each pipeline, we also calculated the permutation feature importance using 100 permutations. In the MDD subgroup with random splitting, the most important feature was reported mood, with the missingness of the mood EMA as the second most important feature. The number of places visited, percent home, age, GPS missingness, platform and sleep were other notable features. While different in exact importance score, the most important features where highly similar among estimators. With the group split GPS missingness was the most important feature with mood EMA, number of places visited, percent home and missingness of the mood EMA as other important predictors. However, these should be interpreted with caution as the different estimators did not perform much better than the Dummy estimators.

**Fig. 1.**
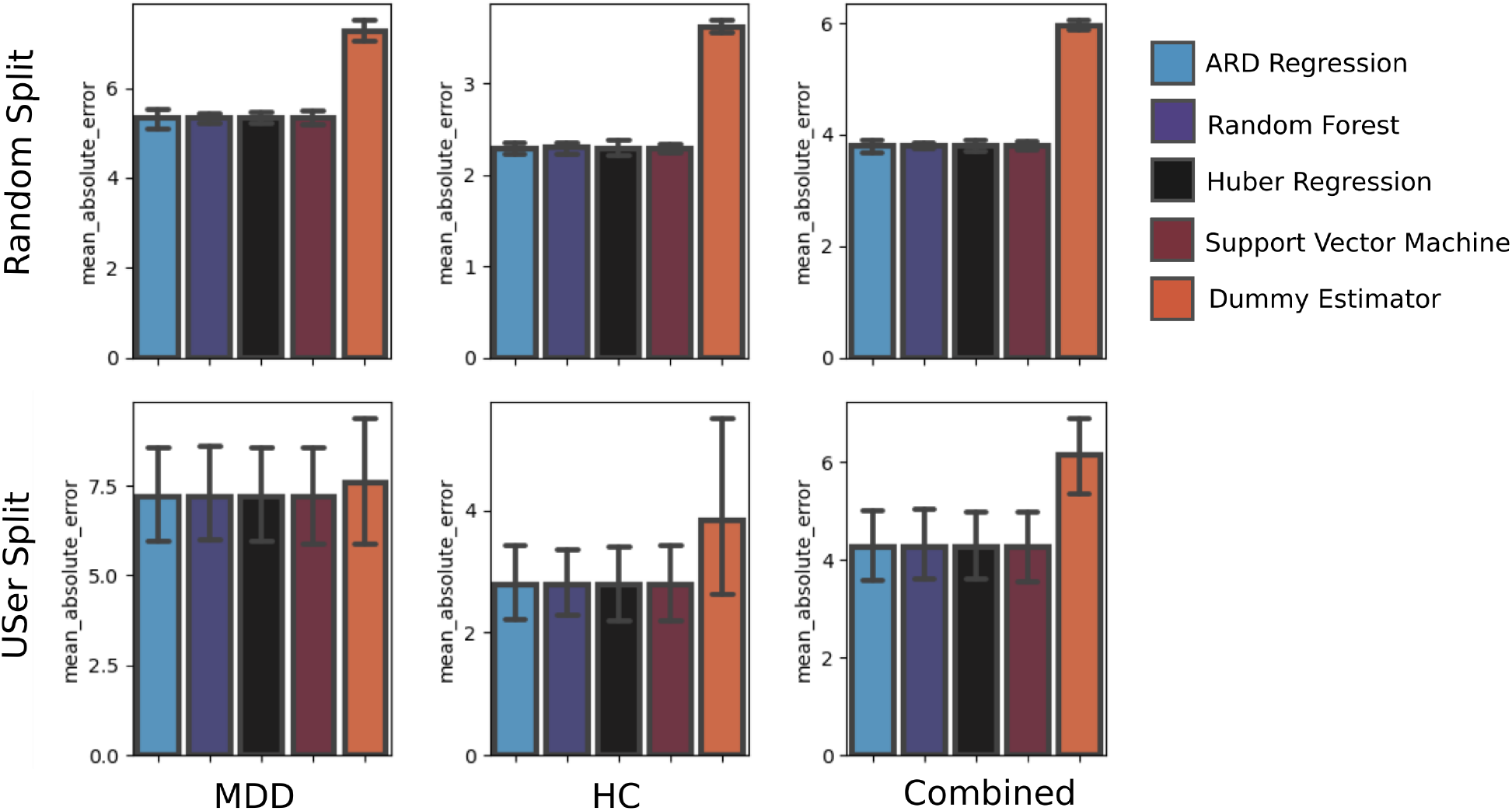
Predicting BDI. Predicting BDI in a random split scenario, all estimators performed better than the dummy estimator that was predicting the average BDI score, with very similar mean absolute errors between the different estimators. The random forest performed best for the MDD subgroup (MAE: 5.316, Pearson’s R: 0.637, Permutation p-value: p<0.001) and Combined subgroup (MAE: 3.791, Pearson’s R: 0.704) while the ARD Regression performed best in the HC subgroup (MAE: 2.280, Pearson’s R: 0.753, Permutation p-value: p<0.001). In the user split scenario, the estimators did not perform much better than the dummy estimator. The best estimator in the group split scenario was the random forest for the MDD (MAE: 7.183, Pearson’s R: 0.475, Permutation p-value: p<0.001), HC (MAE: 2.786, Pearson’s R: 0.383, Permutation p-value: p<0.001) and Combined (MAE: 4.259, Pearson’s R: 0.610, Permutation p-value: p<0.001) subgroups. The performance here should be interpreted with caution however as no estimator strongly outperformed the dummy estimator. *MAE: Mean absolute error.

In the HC subgroup with random splitting, the most important feature was the mood EMA followed by missingness of the mood EMA, sex and age. Again, feature importances were highly similar among estimators. With user splitting the most important feature was number of places visited, followed by mood EMA missingness, percent home, GPS missingness, age and sex. Again these need to be interpreted with caution as the estimators did not perform much better than the Dummy estimator.

In the combined group with random splitting, the most important feature was the mood EMA, followed by GPS missingness, mood EMA missingness, sex and number of places visited. With user splitting the most important feature was mood EMA followed by GPS missingness, mood EMA missingness percent home and age. Within both splitting scenarios, there was a high degree of similarity between the feature importances for the different estimators.

### BDI correlates with decreased mobility

With GPS-derived mobility measures as being some of the important features in our predictive models we next looked at the association between different geolocation-derived metrics and BDI scores. This was done in an exploratory manner to provide possible explanations for the observed performance.

We predicted BDI using either percent home or number of places visited with age and sex as covariates in a linear mixed model with a variable intercept for each subject. We also ran each model separately for the depressed (MDD) and healthy (HC) subgroups to account for differences in trends when analyzing the different subgroups compared to analyzing all subjects together, also known as Simpson’s paradox [31]. When predicting BDI using percent home in the MDD subgroup, neither percent home (p=0.796, z=0.259), age (p=0.767, z=0.297) nor sex (p=0.793, z=-0.262) were significant predictors of BDI. In the HC subgroup percent home was a significant predictor (p<0.001, z=3.958), while age (p=0.405, z=0.833) and sex (p=0.293, z=1.051) were not. When predicting BDI using number of places visited in the MDD subgroup, the number of places (p=0.003, z=3.016) was a significant predictor, while age (p=0.758, z=0.308) and sex (p=0.788, z=-0.269) were not. In the HC subgroup neither number of places (p=0.171, z=-1.369), age (p=0.383, z=0.872) nor sex (p=0.253, z=1.142) was significant.

### BDI correlates with worsened mood and sleep

As the mood EMA was one of the strongest predictors in all models with good predictive performance, we investigated the utility of self-reported measures of mood and sleep (ecological momentary assessments, EMAs) by using them as predictors of BDI. As before we ran two separate regressions for HC and MDD to account for a potential Simpson’s paradox [31].

**Fig. 2.**
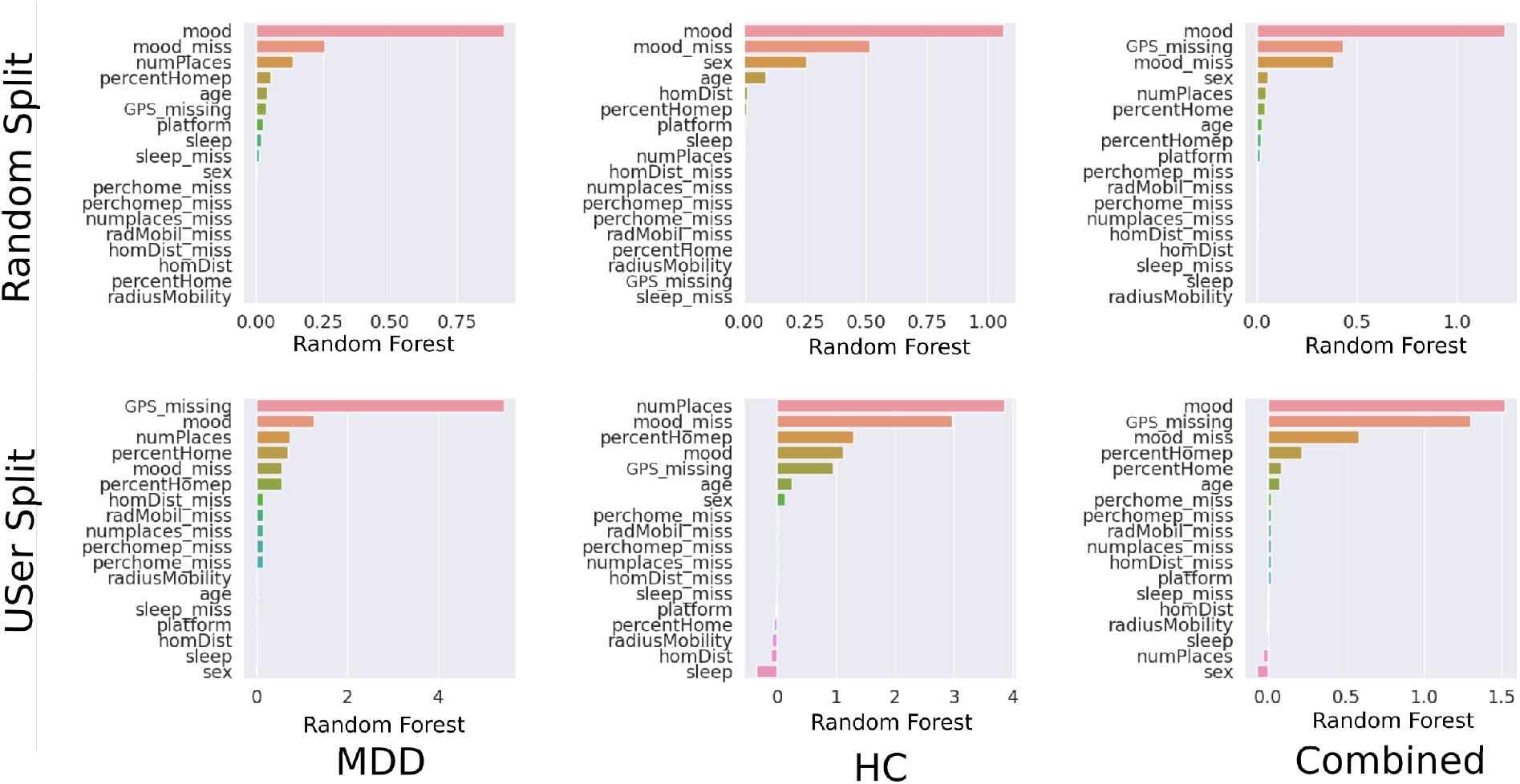
Permutation Feature Importance. We calculated the permutation feature importance for every pipeline. Mood EMA emerged as the most important predictor in the random split scenario for every pipeline. Other important features were the missingness of the mood EMA, the missingness of the GPS data, the number of places visited, the percent home, age and sex. For the user split the pipelines did not perform much better than the Dummy estimator for the MDD subgroup therefore interpreting the feature importance is not encouraged. In the combined subgroup where the pipelines outperformed the Dummy estimator mood again emerged as the most important feature with GPS missingness, mood EMA missingness, percent home and age as additional important features. *Mood: Mood EMA, sleep: Sleep EMA, numPlaces: Number of places visited, percentHome: Percent home, percentHomep: normalised percent Home, homDist: Distance from home, radiusMobility: Radius of Mobility, age: age, sex: Sex, platform: phone platform, GPS_missing: missingness of GPS data, mood_miss: Missingness of mood EMA, sleep_miss: Missingness of sleep EMA, percenthome_miss: Missingness of percent home, percenthomep_miss: Missingness of normalised percent home, numplaces_miss: Missingness of number of places, radMobil_miss: Missingness of Radius of Mobility, homDist_miss: Missingness of radius of Mobility

For predicting BDI from self-reported mood we used 3753 data points from 132 subjects (53 MDD, 79 HC). We used a linear mixed model as described above with age and sex as covariates. For the MDD subgroup, we found a significant effect of mood (p<0.001, z=-22.612), but not age (p=0.931, z=0.087) or sex (p=0.943, z=0.071). In the HC subgroup, we again found a significant effect of mood (p<0.001, z=-23.653), but not age (p=0.343, z=0.947) or sex (p=0.345, z=0.730).

For predicting BDI from self-reported sleep we used 3781 data points from 132 subjects (54 MDD, 78 HC). For the MDD subgroup, we found a significant effect of self-reported sleep (p<0.001, z=4.267), but not age (p=0.739, z=0.333) or sex (p=0.782, z=-0.276). For the HC subgroup, we found a significant effect of self-reported sleep (p=0.004, z=-2.912), but not age (p=0.448, z=0.759) or sex (p=0.278, z=1.084).

### Diagnosis correlates with mobility markers

After establishing the relation between BDI and geolocation markers of mobility we looked at the association between GPS-derived mobility markers and diagnosis. We extracted the average percent home and average number of places visited for each healthy control and each subject with a diagnosis of major depressive disorder. 156 individuals (60 MDD, 96 HC) were included in the analysis (see Table 16). We predicted one of the two mobility measures, average mood EMA response or average sleep EMA response with diagnosis, age and sex as predictors using a linear regression model.

Predicting percent home we saw a significant effect of sex (p<0.001, t=3.621), diagnosis (p=0.009, t=2.633) and age (p=0.018, t=2.389). A diagnosis of MDD was associated with an increased amount of time spent home (coeff=1.538, CI=0.384 - 2.692). Predicting the number of places visited we saw a significant effect of sex (p=0.016, t=-2.429), diagnosis (p=0.010, t=-2.593) and age (p=0.019, t=-2.376). A diagnosis of MDD was associated with a decreased number of places visited (coeff=-0.552, CI=-0.972 - -0.0131). When predicting average mood we found a strongly significant effect for diagnosis (p<0.001, t=-6.869) where a diagnosis of MDD was negatively correlated with (coeff=-1.537, CI=-1.98 - -1.095), without a significant effect for age (p=0.645, t=-0.461) or sex (p=0.816, t=-0.233). For average reported sleep we found no significant effect of diagnosis (p=0.897, t=-0.130). The effects for diagnosis on number of places, percent home and average mood remained significant even after adjusting for multiple comparisons using the Bonferroni method.

## Discussion

This study aimed to explore the feasibility of predicting symptom scores from smartphone data, focusing on an area that had received limited attention in previous research, compared to classification. Unlike earlier models that primarily concentrated on predicting item-level symptoms or overall diagnosis state, our analysis focused on predicting symptom scores using different estimators. Most previous studies used machine learning models to classify patients based on their phone data as depressed or healthy [11, 17, 20]. Other studies had demonstrated success in predicting specific aspects of depression but still used classifiers to predict individual symptom-level items, rather than predicting composite scores that rate depression severity [32, 33]. Notably some studies predicted depression severity using composite scores, using psychometric instruments other than BDI. These studies also did not explicitly model missingness as they mostly relied on passive features. Here our study adds to a growing body of evidence for the utility of monitoring depressive symptomatology.

**Fig. 3.**
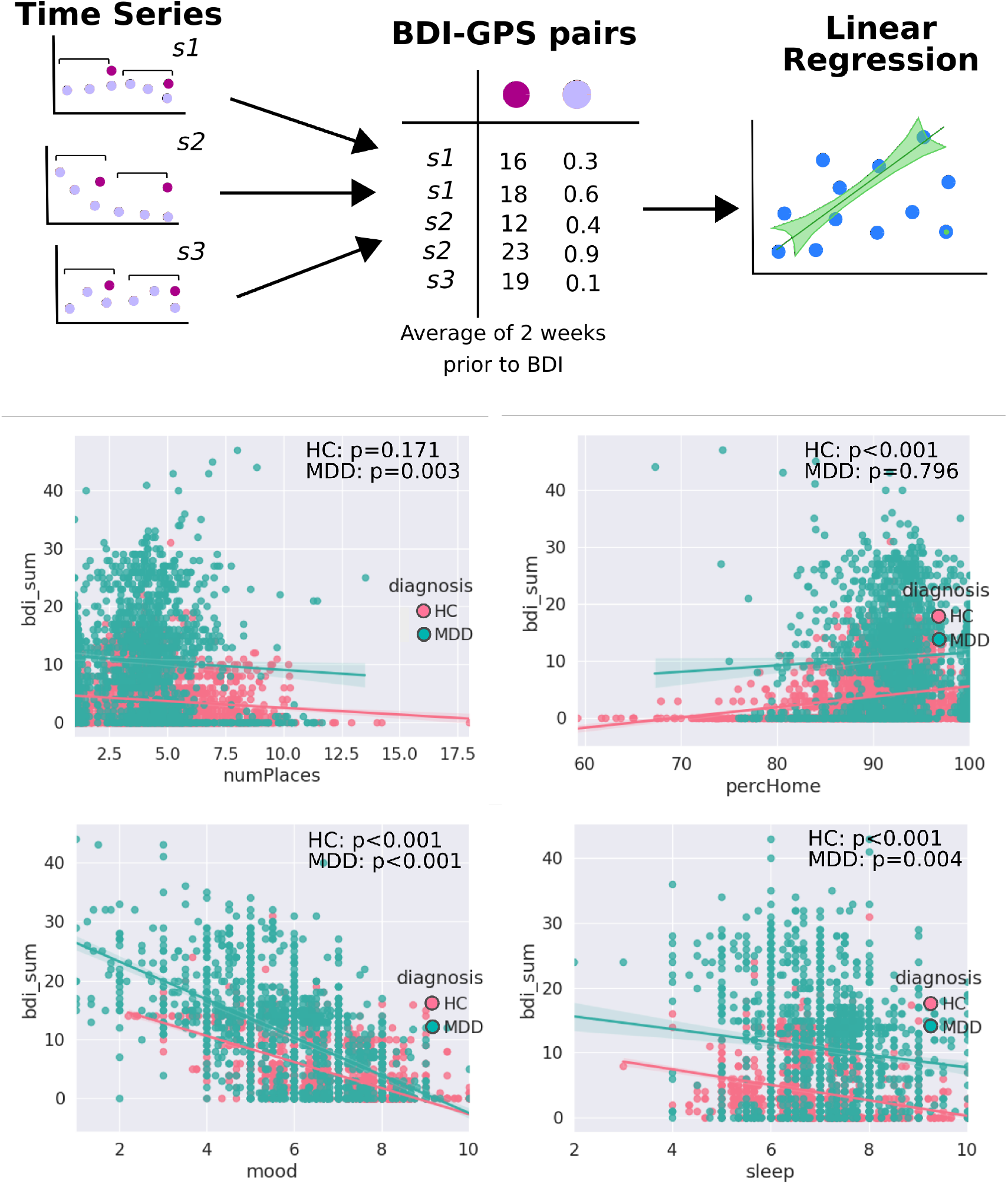
Predictors of BDI. GPS-derived mobility markers are partially significant predictors in the MDD subgroup (number of places: p=0.003, percent home: p=0.796) as well as the HC subgroup (number of places: p=0.171, percent home: p<0.001)BDI is also significantly associated with mood (p<0.001), sleep (p=0.004) in the MDD subgroup as well as the HC subgroup (mood: p<0.001; sleep: p<0.001).

Our results show that employed models outperformed the Dummy baseline estimator in a random split scenario, albeit with only a marginal improvement in a user split scenario. This is in line with previous findings by Lewis et al, which found that classifiers work better than baseline estimators in random and time-split scenarios, but not in user-split scenarios [24]. First, these models should be evaluated in larger, more heterogeneous samples, as the relatively small dataset used in the study may have impacted the models’ performance. Larger and more diverse samples could yield better results. Secondly, models should be trained in cohorts where individuals show greater variability of symptom ranges enabling models to leverage more meaningful within-person variation. Third, for clinical deployment, it might be necessary to collect a few data points from a user and then adjust the model based on this user’s data. To further improve prediction accuracy, model personalization using mixed effects machine learning models, as proposed by Lewis et al., could be considered [24].

Among the features analyzed using permutation feature importance, the mood ecological momentary assessment (EMA) emerged as the most influential predictor, backed by highly significant effects in our mixed models, followed by the missingness of different signals and GPS-derived mobility features. The assessment of feature importance was only meaningful in the random split scenario as in the user-split scenarios models failed to significantly predict mood above baseline. In this scenario interpreting feature importances should not be attempted. For future studies, it might be important to effectively model missingness as it may provide crucial information for machine learning models to achieve accurate predictions.

**Fig. 4.**
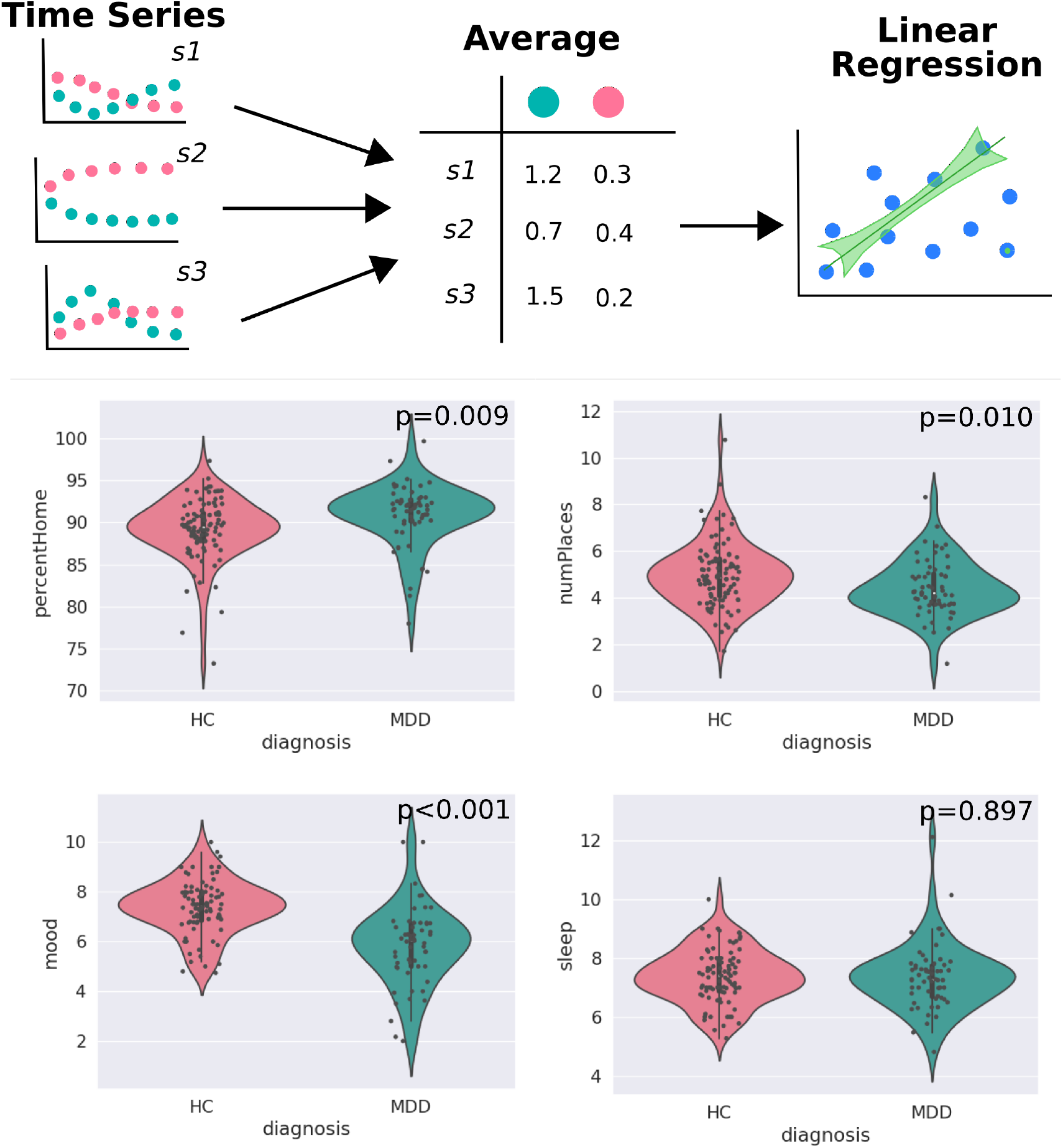
Diagnosis predicts lower mood, more time at home and decreased number of places. When using diagnosis as a predictor for GPS-derived mobility features we find a significantly increased percent home (p=0.009) and a significantly decreased number of places visited (p=0.010). A diagnosis of depression also predicts a significantly lower average mood (p<0.001). We do not find evidence for a significant difference in average self-reported sleep quality (p=0.897).

In our investigation using mixed modelling to explore the connection between distinct predictors and BDI scores, we confirmed the robust predictive capability of mood EMAs, while also establishing significant effects for GPS-derived mobility markers. These findings reinforce the strength of EMAs as a key indicator of depression severity. Previous research has consistently demonstrated substantial links between EMA scores and depression scales within psychiatric populations and other study groups [15, 34]. The empirical evidence for GPS-derived mobility markers also aligns with existing studies, which have highlighted the correlation between alterations in these markers and shifts in depressive symptoms [35]. Despite notable levels of missing data in our GPS records, we replicate effects from prior investigations. Enhancing the frequency and reliability of data sampling could potentially yield more robust and meaningful estimates. However, it’s important to note that while EMAs exhibit effectiveness, their reliance on active user participation may limit their widespread adoption in certain contexts [34], thus impacting their clinical utility.

Considering the group-level findings, we show that depression is significantly associated with the average reported mood and GPS-derived mobility markers. Both number of places and percent home have previously been shown to be significantly associated with a diagnosis of depression across multiple studies [11, 17]. The average sleep quality has also been previously associated with depression [36–38], yet we are unable to reproduce this finding. While this might be due to depressed patients experiencing different levels of symptom severity being ignored when examining the average, self-reported sleep did not emerge as a strong predictor of BDI score in the feature importance analysis. Overall our group-level analysis is in line with findings from previous studies.

While predicting composite scores is an important task in remote monitoring, it may not fully capture the variability in patient symptoms. Depression is considered a highly heterogeneous disease, with some authors suggesting it might be multiple diseases disguised under a common symptom. Therefore, to gain a more comprehensive overview of a patient’s condition, predicting individual items or symptom groups such as in latent factor models might provide a more holistic view of depression severity and is potentially easier to predict as these categories are better reflected in measured outcomes than composite scores. These factor models have also been shown to track multiple dimensions of psychiatric illness as well as general psychopathology [39].

Other challenges for clinical utility concern participant dropout, especially for models that rely on active features such as EMAs. To address this future apps will need to be carefully designed to become more engaging and promote user adherence, without strongly altering user behaviour. Models derived exclusively from passive sensing are less prone to these problems as they do not rely on active user participation. Nonetheless, the lack of standardization among sensors across various studies and applications introduces challenges in the transference of models. Sensor harmonization, a pivotal challenge, is influenced by factors such as the diversity in the count and categories of phone sensors captured in different studies. In this context, our study offers valuable insights into the array of phone-derived metrics that hold significance for predicting depression severity, contributing to a deeper understanding of this challenge.

Overall, this study contributes valuable evidence to the potential utility of smartphone data in mood state monitoring. However, future studies should explore the prospect of predicting depression further ahead in time, potentially serving as an early-warning tool for better intervention and management of the condition. Additionally, addressing the challenges and limitations identified in this study could pave the way for more robust and effective depression monitoring solutions using smartphone-based data.

## Data Availability

As data was derived from multiple study protocols with different limitations, the potential sharing of data would need to be discussed and evaluated on a case-by-case basis with the senior authors and different study leaders

## ACKNOWLEDGEMENTS

Funding was in part provided by grants from the German Science Foundation (DFG): Project number 44541416-TRR 58 (CRC-TRR58, Projects C09 and Z02 to Udo Dannlowski). Further DFG funding as part of Forschungsgruppe/Research Unit FOR2107 was received by: Tilo Kircher (speaker FOR2107; DFG grant numbers KI 588/14-1, KI 588/14-2, KI 588/15-1, KI 588/17-1), Udo Dannlowski (co-speaker FOR2107; DA 1151/5-1, DA 1151/5-2, DA 1151/6-1), Axel Krug (KR 3822/5-1, KR 3822/7-2), Igor Nenadic (NE 2254/1-2, NE 2254/2-1, NE 2254/3-1, NE 2254/4-1), Carsten Konrad (KO 4291/3-1), Marcella Rietschel (RI 908/11-1, RI 908/11-2), Markus Noethen (NO 246/10-1, NO 246/10-2), Stephanie Witt (WI 3439/3-1, WI 3439/3-2), Andreas Jansen (JA 1890/7-1, JA 1890/7-2), Tim Hahn (HA 7070/2-2, HA7070/3, HA7070/4), Bertram Mueller-Myhsok (MU1315/8-2), Astrid Dempfle (DE 1614/3-1, DE 1614/3-2), Petra Pfefferle (PF 784/1-1, PF 784/1-2), Harald Renz (RE 737/20-1, 737/20-2), Carsten Konrad (KO 4291/4-1).

Interdisciplinary Center for Clinical Research (IZKF) of the medical faculty of Muenster provided funding to Udo Dannlowski (Grant Dan3/012/17) and Nils Opel (SEED 11/19)

“Innovative Medizinische Forschung” (IMF) of the medical faculty of Münster provided funding to Elisabeth J. Leehr (Grant LE121703 and LE121904), Nils Opel and Tim Hahn (Grant OP121710).

## Supplementary Material

### A. ReMAP App

The Remote Monitoring Application in Psychiatry (ReMAP) app was developed by researchers from the Department of Psychiatry at the University of Münster. It enables high-resolution monitoring of activity using smartphone sensors, including geolocation, steps, walking distance, and acceleration. The app also incorporates active self-reports on sleep, mood, and voice samples. Every two weeks, participants were given the option to complete the Beck Depression Inventory (BDI) with a random variance of two days. Participation in the self-reporting was voluntary, and it was not a requirement for financial compensation. ReMAP was used as an additional assessment tool for participants in various ongoing longitudinal studies, primarily focusing on neuroimaging studies involving structural and functional MRI assessments, genotyping, and the evaluation of various clinical variables.

### B. ReMAP Participants

The sample for the analyses in this paper was drawn from several ongoing longitudinal cohorts. Included samples stem from the Marburg/Münster Affective Disorder Cohort Study (MACS, n=95) (Vogelbacher et al., 2018), the Münster Neuroimage Cohort (MNC, n=27) (Dannlowski et al., 2016; Opel et al., 2019), two subsamples of the SFB-TRR58 cohort (n=23; Z02 Münster cohort and SpiderVR Münster cohort (Schwarzmeier et al., 2020)), the TIP (n=6) cohort and the SEED cohort (n=5). All cohorts comprise healthy control (HC) participants, as well as different patient groups. Both, the MACS and the MNC cohorts include major depressive disorder (MDD) and bipolar disorder (BD) patients under current or former inpatient treatment. The SFB-TRR58 sample includes patients with a spider phobia (SP). The TIP sample includes patients with a social anxiety disorder (SAD), MDD, or comorbid SAD and MDD. For our analyses, we excluded all patients that did not meet the criteria for healthy controls or major depressive disorder.

### C. Predictive Modelling

Results for all the different estimator pipelines can be found in table 4. Condition refers to the subgroup the pipeline was trained on, while split refers to the splitting procedure for the data. Tables 2 and 3 provide information on the HC and MDD subgroups used in the predictive modelling.

**Table 1.** Demographic Summary of all subjects used for predicting BDI using machine learning models. For further information on subgroups see appendix.

| Demographic variable | Value |
| --- | --- |
| Age range | 18 - 67 |
| Age median | 35 ± 14.04 |
| Sex ratio | 103 female, 39 male |
| MDD-HC ratio | 57 MDD, 85 HC |
| BDI range | 0 - 47 |

**Table 2.** Demographic Summary for the MDD subgroup of the BDI prediction model.

| Demographic variable | Value |
| --- | --- |
| Age range | 19-62 |
| Age median | 37.0 ± 12.61 |
| Sex ratio | 42 female, 15 male |
| BDI range | 0-47 |
| BDI median | 7 ± 8.75 |

**Table 3.** Demographic Summary for the HC subgroup of the BDI prediction model.

| Demographic variable | Value |
| --- | --- |
| Age range | 18-67 |
| Age median | 35.0 ± 15.00 |
| Sex ration | 61 female, 24 male |
| BDI range | 0-31 |
| BDI median | 2 ± 4.56 |

**Table 4.** Results for the different classifiers trained on the MDD cohort. Condition refers to the subgroup, split to the data splitting and estimator to the estimator used in the pipeline. *MAE: Mean absolute error; MSE: Mean squared error; Spearman: Spearman Rank Correlation; Pearson: Pearson correlation; r-pval: p-value of the permutation test for the correlation between predicted and true values of the target variable.

| Condition | Split | Estimator | MAE | MSE | Spearman | Pearson | r-pval |
| --- | --- | --- | --- | --- | --- | --- | --- |
| Comb | User | ARD | 4.261 | 32.976 | 0.549 | 0.608 | <0.001 |
| Comb | User | Dummy | 6.14 | 56.842 | 0 | 0 | 0 |
| Comb | User | HuberRegressor | 4.260 | 32.977 | 0.550 | 0.607 | <0.001 |
| Comb | User | RandomForest | 4.259 | 32.929 | 0.550 | 0.610 | <0.001 |
| Comb | User | SVR | 4.260 | 32.980 | 0.549 | 0.609 | <0.001 |
| Comb | Random | ARD | 3.794 | 27.070 | 0.713 | 0.701 | <0.001 |
| Comb | Random | Dummy | 5.962 | 54.033 | 0 | 0 | 0 |
| Comb | Random | HuberRegressor | 3.798 | 27.122 | 0.712 | 0.706 | <0.001 |
| Comb | Random | RandomForest | 3.791 | 27.064 | 0.713 | 0.704 | <0.001 |
| Comb | Random | SVR | 3.797 | 27.112 | 0.713 | 0.706 | <0.001 |
| HC | User | ARD | 2.787 | 12.163 | 0.353 | 0.384 | <0.001 |
| HC | User | Dummy | 3.823 | 23.598 | 0 | 0 | 0 |
| HC | User | HuberRegressor | 2.787 | 12.163 | 0.353 | 0.384 | <0.001 |
| HC | User | RandomForest | 2.786 | 12.160 | 0.353 | 0.383 | <0.001 |
| HC | User | SVR | 2.787 | 12.162 | 0.353 | 0.384 | <0.001 |
| HC | Random | ARD | 2.280 | 9.024 | 0.678 | 0.753 | <0.001 |
| HC | Random | Dummy | 3.620 | 20.845 | 0 | 0 | 0 |
| HC | Random | HuberRegressor | 2.289 | 9.102 | 0.676 | 0.752 | <0.001 |
| HC | Random | RandomForest | 2.288 | 9.094 | 0.677 | 0.749 | <0.001 |
| HC | Random | SVR | 2.286 | 9.080 | 0.678 | 0.751 | <0.001 |
| MDD | User | ARD | 7.183 | 74.887 | 0.373 | 0.475 | <0.001 |
| MDD | User | Dummy | 7.591 | 82.810 | 0 | 0 | 0 |
| MDD | User | HuberRegressor | 7.183 | 74.887 | 0.373 | 0.475 | <0.001 |
| MDD | User | RandomForest | 7.183 | 74.874 | 0.374 | 0.475 | <0.001 |
| MDD | User | SVR | 7.184 | 74.886 | 0.374 | 0.473 | <0.001 |
| MDD | Random | ARD | 5.324 | 45.026 | 0.679 | 0.645 | <0.001 |
| MDD | Random | Dummy | 7.258 | 76.661 | 0 | 0 | 0 |
| MDD | Random | HuberRegressor | 5.327 | 45.109 | 0.677 | 0.643 | <0.001 |
| MDD | Random | RandomForest | 5.316 | 45.344 | 0.677 | 0.637 | <0.001 |
| MDD | Random | SVR | 5.322 | 44.895 | 0.678 | 0.644 | <0.001 |

### D. Mixed Modelling of BDI

The demographics and results for the different regression models on BDI can be seen in tables 5 to 15. The formula for the linear mixed models can be read as *BDI ∼ var* + *age* + *sex* + (1 | *subject*) where var stands for the variable of interest such as mood EMA, sleep EMA, percent home or the number of places visited.

**Table 5.** Demographic Summary for estimating the effect of GPS-derived mobility markers on BDI (N=140, 4011 data points).

| Demographic variable | Value |
| --- | --- |
| Age range | 18-67 |
| Age median | 35.5 ± 14.09 |
| Sex ratio | 101 female, 39 male |
| MDD-HC ratio | 56 MDD, 84 HC |

**Table 6.** Model summary for correlating BDI with percent home in MDD.

|  | Coef. | Std.Err. | z | P>\|z\| | [0.025 | 0.975] |
| --- | --- | --- | --- | --- | --- | --- |
| Intercept | 10.525 | 5.874 | 1.792 | 0.073 | -0.988 | 22.037 |
| sex[T.w] | -0.795 | 3.030 | -0.262 | 0.793 | -6.734 | 5.145 |
| age | 0.032 | 0.107 | 0.297 | 0.767 | -0.178 | 0.242 |
| percHome | 0.007 | 0.027 | 0.259 | 0.796 | -0.045 | 0.059 |
| userId Var | 89.596 | 5.015 |  |  |  |  |

**Table 7.** Model summary for correlating BDI with percent home in HC.

|  | Coef. | Std.Err. | z | P>\|z\| | [0.025 | 0.975] |
| --- | --- | --- | --- | --- | --- | --- |
| Intercept | -2.092 | 1.549 | -1.351 | 0.177 | -5.128 | 0.943 |
| sex[T.w] | 0.870 | 0.828 | 1.051 | 0.293 | -0.752 | 2.493 |
| age | 0.021 | 0.025 | 0.833 | 0.405 | -0.028 | 0.070 |
| percHome | 0.036 | 0.009 | 3.958 | 0.000 | 0.018 | 0.054 |
| userId Var | 9.844 | 0.772 |  |  |  |  |

**Table 8.** Model summary for correlating BDI with number of places in MDD.

|  | Coef. | Std.Err. | z | P>\|z\| | [0.025 | 0.975] |
| --- | --- | --- | --- | --- | --- | --- |
| Intercept | 10.372 | 5.376 | 1.929 | 0.054 | -0.164 | 20.908 |
| sex[T.w] | -0.812 | 3.020 | -0.269 | 0.788 | -6.731 | 5.107 |
| age | 0.033 | 0.107 | 0.308 | 0.758 | -0.177 | 0.243 |
| numPlaces | 0.193 | 0.064 | 3.016 | 0.003 | 0.068 | 0.319 |
| userId Var | 89.000 | 4.995 |  |  |  |  |

**Table 9.**
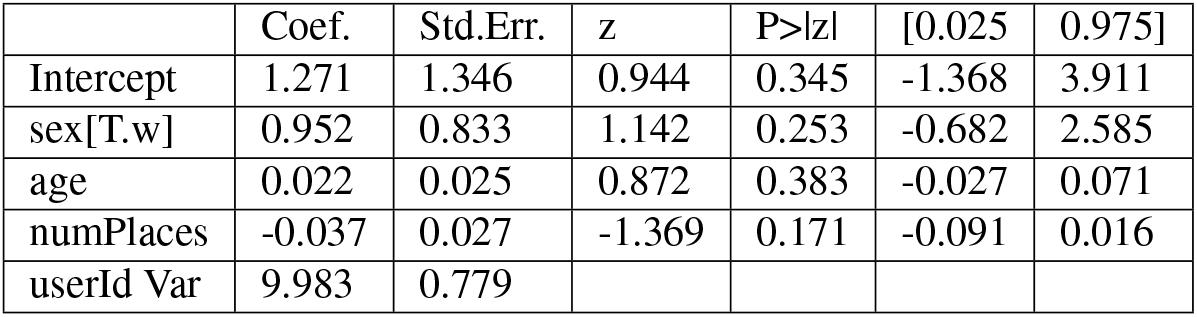
Model summary for correlating BDI with number of places in HC.

|  | Coef. | Std.Err. | z | P>\|z\| | [0.025 | 0.975] |
| --- | --- | --- | --- | --- | --- | --- |
| Intercept | 1.271 | 1.346 | 0.944 | 0.345 | -1.368 | 3.911 |
| sex[T.w] | 0.952 | 0.833 | 1.142 | 0.253 | -0.682 | 2.585 |
| age | 0.022 | 0.025 | 0.872 | 0.383 | -0.027 | 0.071 |
| numPlaces | -0.037 | 0.027 | -1.369 | 0.171 | -0.091 | 0.016 |
| userId Var | 9.983 | 0.779 |  |  |  |  |

**Table 10.** Demographic Summary for estimating the effect of self-reported mood on BDI (N=132, 3753 data points).

| Demographic variable | Value |
| --- | --- |
| Age range | 18-67 |
| Age median | 36.5 ± 14.17 |
| Sex ration | 97 female, 35 male |
| MDD-HC ratio | 53 MDD, 79 HC |

**Table 11.** Model summary for correlating BDI with self-reported mood in MDD.

|  | Coef. | Std.Err. | z | P>\|z\| | [0.025 | 0.975] |
| --- | --- | --- | --- | --- | --- | --- |
| Intercept | 20.763 | 4.889 | 4.247 | 0.000 | 11.180 | 30.346 |
| sex[T.w] | 0.197 | 2.761 | 0.071 | 0.943 | -5.214 | 5.607 |
| age | 0.008 | 0.094 | 0.087 | 0.931 | -0.177 | 0.193 |
| mood | -1.619 | 0.072 | -22.612 | 0.000 | -1.759 | -1.479 |
| userId Var | 66.379 | 4.425 |  |  |  |  |

**Table 12.** Model summary for correlating BDI with self-reported mood in HC.

|  | Coef. | Std.Err. | z | P>\|z\| | [0.025 | 0.975] |
| --- | --- | --- | --- | --- | --- | --- |
| Intercept | 9.336 | 1.191 | 7.841 | 0.000 | 7.002 | 11.670 |
| sex[T.w] | 0.247 | 0.714 | 0.345 | 0.730 | -1.153 | 1.647 |
| age | 0.020 | 0.021 | 0.947 | 0.343 | -0.022 | 0.062 |
| mood | -1.047 | 0.044 | -23.653 | 0.000 | -1.134 | -0.961 |
| userId Var | 6.784 | 0.634 |  |  |  |  |

**Table 13.** Demographic Summary for estimating the effect of sleep on BDI (N=132, 3781 data points).

| Demographic variable | Value |
| --- | --- |
| Age range | 18-67 |
| Age median | 36.5 ± 14.17 |
| Sex ration | 97 female, 35 male |
| MDD-HC ratio | 54 MDD, 78 HC |

**Table 14.**
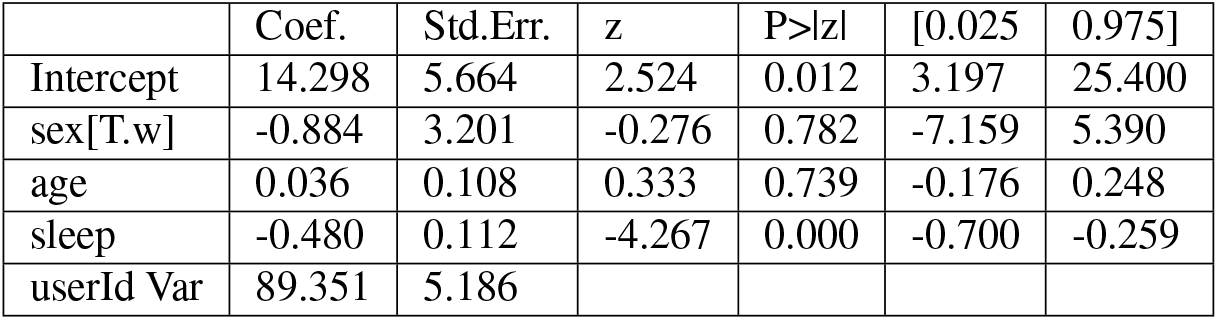
Model summary for correlating BDI with self-reported sleep in MDD.

|  | Coef. | Std.Err. | z | P>\|z\| | [0.025 | 0.975] |
| --- | --- | --- | --- | --- | --- | --- |
| Intercept | 14.298 | 5.664 | 2.524 | 0.012 | 3.197 | 25.400 |
| sex[T.w] | -0.884 | 3.201 | -0.276 | 0.782 | -7.159 | 5.390 |
| age | 0.036 | 0.108 | 0.333 | 0.739 | -0.176 | 0.248 |
| sleep | -0.480 | 0.112 | -4.267 | 0.000 | -0.700 | -0.259 |
| userId Var | 89.351 | 5.186 |  |  |  |  |

**Table 15.**
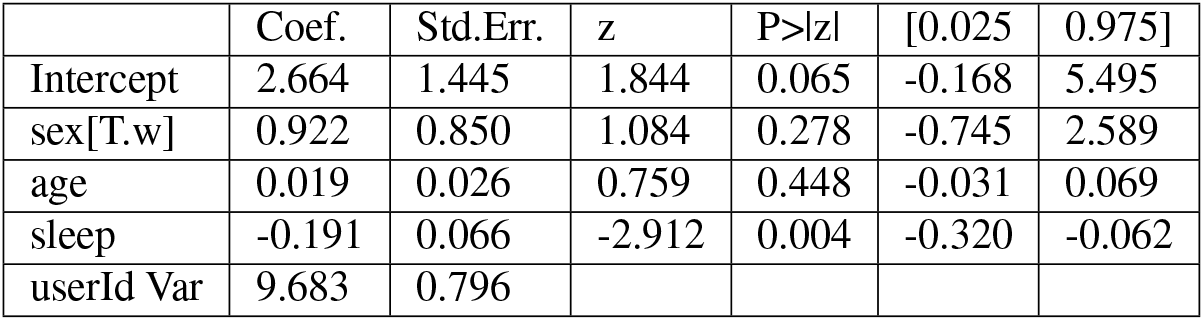
Model summary for correlating BDI with self-reported sleep in HC.

|  | Coef. | Std.Err. | z | P>\|z\| | [0.025 | 0.975] |
| --- | --- | --- | --- | --- | --- | --- |
| Intercept | 2.664 | 1.445 | 1.844 | 0.065 | -0.168 | 5.495 |
| sex[T.w] | 0.922 | 0.850 | 1.084 | 0.278 | -0.745 | 2.589 |
| age | 0.019 | 0.026 | 0.759 | 0.448 | -0.031 | 0.069 |
| sleep | -0.191 | 0.066 | -2.912 | 0.004 | -0.320 | -0.062 |
| userId Var | 9.683 | 0.796 |  |  |  |  |

**Table 16.** Demographic Summary for estimating the effect of diagnosis on GPS-derived mobility markers (N=156).

| Demographic variable | Value |
| --- | --- |
| Age range | 18-67 |
| Age median | 33 ± 13.89 |
| Sex ration | 113 female, 43 male |
| MDD-HC ratio | 60 MDD, 96 HC |

### E. Linear model for diagnosis effects

Tables 16 to 20 provide information on the linear models used to investigate diagnosis effects. The formula for the linear models can be read as *var ∼ diagnosis* + *age* + *sex*, with var standing for the variable of interest. Note that the the average for the variable of interest is used in these models.

**Table 17.** Model Summary for correlating Percent Home with Diagnosis.

|  | coef | std err | t | P>\|t\| | [0.025 | 0.975] |
| --- | --- | --- | --- | --- | --- | --- |
| Intercept | 85.6205 | 1.113 | 76.896 | 0.0 | 83.421 | 87.82 |
| sex[female] | 2.4234 | 0.669 | 3.621 | 0.0 | 1.101 | 3.746 |
| diagnosis[MDD] | 1.5383 | 0.584 | 2.633 | 0.009 | 0.384 | 2.692 |
| age | 0.0516 | 0.022 | 2.389 | 0.018 | 0.009 | 0.094 |

**Table 18.** Model Summary for correlation Number of Places with diagnosis.

|  | coef | std err | t | P>\|t\| | [0.025 | 0.975] |
| --- | --- | --- | --- | --- | --- | --- |
| Intercept | 6.119 | 0.406 | 15.088 | 0.0 | 5.318 | 6.92 |
| sex[T.w] | -0.5921 | 0.244 | -2.429 | 0.016 | -1.074 | -0.111 |
| diagnosis[T.MDD] | -0.5517 | 0.213 | -2.593 | 0.01 | -0.972 | -0.131 |
| age | -0.0187 | 0.008 | -2.376 | 0.019 | -0.034 | -0.003 |

**Table 19.** Model Summary for correlating Mood with Diagnosis.

|  | coef | std err | t | P>\|t\| | [0.025 | 0.975] |
| --- | --- | --- | --- | --- | --- | --- |
| Intercept | 7.5547 | 0.43 | 17.572 | 0.0 | 6.705 | 8.405 |
| sex[T.w] | -0.0603 | 0.259 | -0.233 | 0.816 | -0.573 | 0.452 |
| diagnosis[T.MDD] | -1.5372 | 0.224 | -6.869 | 0.0 | -1.98 | -1.095 |
| age | -0.0038 | 0.008 | -0.461 | 0.645 | -0.02 | 0.012 |

**Table 20.** Model Summary for correlating self-reported sleep with Diagnosis.

|  | coef | std err | t | P>\|t\| | [0.025 | 0.975] |
| --- | --- | --- | --- | --- | --- | --- |
| Intercept | 7.8337 | 0.328 | 23.899 | 0.0 | 7.186 | 8.482 |
| sex[T.w] | -0.1227 | 0.197 | -0.625 | 0.533 | -0.511 | 0.266 |
| diagnosis[T.MDD] | -0.0219 | 0.169 | -0.13 | 0.897 | -0.356 | 0.312 |
| age | -0.0098 | 0.006 | -1.566 | 0.12 | -0.022 | 0.003 |

## Bibliography

1. Deborah S. Hasin, Aaron L. Sarvet, Jacquelyn L. Meyers, Tulshi D. Saha, W. June Ruan, Malka Stohl, and Bridget F. Grant. Epidemiology of adult DSM-5 major depressive disorder and its specifiers in the United States. JAMA Psychiatry, 75(4), 2018. ISSN 2168622X. doi: 10.1001/jamapsychiatry.2017.4602.

2. Jean Pierre Lépine and Mike Briley. The increasing burden of depression. Neuropsychiatric Disease and Treatment, 7(SUPPL.), 2011. ISSN 11782021. doi: 10.2147/NDT.S19617.

3. Harold W. Neighbors, Steven J. Trierweiler, Briggett C. Ford, and Jordana R. Muroff. Racial differences in DSM diagnosis using a semi-structured instrument: The importance of clinical judgment in the diagnosis of African Americans. Journal of Health and Social Behavior, 44 (3), 2003. ISSN 00221465. doi: 10.2307/1519777.

4. Astha Singhal, Yu Yu Tien, and Renee Y. Hsia. Racial-ethnic disparities in opioid prescriptions at emergency department visits for conditions commonly associated with prescription drug abuse. PLoS ONE, 11(8), 2016. ISSN 19326203. doi: 10.1371/journal.pone.0159224.

5. John Torous, Mathew V. Kiang, Jeanette Lorme, and Jukka Pekka Onnela. New tools for new research in psychiatry: A scalable and customizable platform to empower data driven smartphone research. JMIR Mental Health, 3(2), 4 2016. ISSN 23687959. doi: 10.2196/mental.5165.

6. Jukka Pekka Onnela and Scott L. Rauch. Harnessing Smartphone-Based Digital Phenotyping to Enhance Behavioral and Mental Health, 6 2016. ISSN 1740634X.

7. Federica Laricchia. Global smartphone penetration rate as share of population from 2016 to 2022, 3 2023.

8. Petroc Taylor. Smartphone user penetration as share of population in the United States from 2018 to 2025, 1 2023.

9. Florian Ferreri, Alexis Bourla, Charles Siegfried Peretti, Tomoyuki Segawa, Nemat Jaafari, and Stéphane Mouchabac. How new technologies can improve prediction, assessment, and intervention in obsessive-compulsive disorder (e-ocd): Review, 2019. ISSN 23687959.

10. Luigi F. Saccaro, Giulia Amatori, Andrea Cappelli, Raffaele Mazziotti, Liliana Dell’Osso, and Grazia Rutigliano. Portable technologies for digital phenotyping of bipolar disorder: A systematic review, 2021. ISSN 15732517.

11. Valeria De Angel, Serena Lewis, Katie White, Carolin Oetzmann, Daniel Leightley, Emanuela Oprea, Grace Lavelle, Faith Matcham, Alice Pace, David C. Mohr, Richard Dobson, and Matthew Hotopf. Digital health tools for the passive monitoring of depression: a systematic review of methods. npj Digital Medicine, 5(1), 12 2022. ISSN 23986352. doi: 10.1038/s41746-021-00548-8.

12. Ansgar Conrad, Frank H. Wilhelm, Walton T. Roth, David Spiegel, and C. Barr Taylor. Circadian affective, cardiopulmonary, and cortisol variability in depressed and nondepressed individuals at risk for cardiovascular disease. Journal of Psychiatric Research, 42(9), 2008. ISSN 00223956. doi: 10.1016/j.jpsychires.2007.08.003.

13. Heejung Kim, Sung Hee Lee, Sang Eun Lee, Soyun Hong, Hee Jae Kang, and Namhee Kim. Depression prediction by using ecological momentary assessment, actiwatch data, and machine learning: Observational study on older adults living alone. JMIR mHealth and uHealth, 7(10), 2019. ISSN 22915222. doi: 10.2196/14149.

14. Jian Cao, Anh Lan Truong, Sophia Banu, Asim A. Shah, Ashutosh Sabharwal, and Nidal Moukaddam. Tracking and predicting depressive symptoms of adolescents using smartphone-based self-reports, parental evaluations, and passive phone sensor data: Development and usability study. JMIR Mental Health, 7(1), 2020. ISSN 23687959. doi: 10.2196/14045.

15. Ilya Baryshnikov, Talayeh Aledavood, Tom Rosenström, Roope Heikkilä, Richard Darst, Kirsi Riihimäki, Outi Saleva, Jesper Ekelund, and Erkki Isometsä. Relationship between daily rated depression symptom severity and the retrospective self-report on PHQ-9: A prospective ecological momentary assessment study on 80 psychiatric outpatients. Journal of Affective Disorders, 324, 2023. ISSN 15732517. doi: 10.1016/j.jad.2022.12.127.

16. Abhishek Pratap, David C. Atkins, Brenna N. Renn, Michael J. Tanana, Sean D. Mooney, Joaquin A. Anguera, and Patricia A. Areán. The accuracy of passive phone sensors in predicting daily mood. Depression and Anxiety, 36(1), 2019. ISSN 15206394. doi: 10.1002/da.22822.

17. Sohrab Saeb, Mi Zhang, Mary Kwasny, Christopher J. Karr, Konrad Kording, and David C. Mohr. The relationship between clinical, momentary, and sensor-based assessment of depression. In Proceedings of the 2015 9th International Conference on Pervasive Computing Technologies for Healthcare, PervasiveHealth 2015, 2015. doi: 10.4108/icst.pervasivehealth.2015.259034.

18. Sohrab Saeb, Emily G. Lattie, Stephen M. Schueller, Konrad P. Kording, and David C. Mohr. The relationship between mobile phone location sensor data and depressive symptom severity. PeerJ, 2016(9), 2016. ISSN 21678359. doi: 10.7717/peerj.2537.

19. Fabian Wahle, Tobias Kowatsch, Elgar Fleisch, Michael Rufer, and Steffi Weidt. Mobile sensing and support for people with depression: A pilot trial in the wild. JMIR mHealth and uHealth, 4(3), 2016. ISSN 22915222. doi: 10.2196/mhealth.5960.

20. Akane Sano, Sara Taylor, Andrew W. McHill, Andrew J.K. Phillips, Laura K. Barger, Elizabeth Klerman, and Rosalind Picard. Identifying objective physiological markers and modifiable behaviors for self-reported stress and mental health status using wearable sensors and mobile phones: Observational study. Journal of Medical Internet Research, 20(6), 2018. ISSN 14388871. doi: 10.2196/jmir.9410.

21. Rui Wang, Weichen Wang, Alex daSilva, Jeremy F. Huckins, William M. Kelley, Todd F. Heatherton, and Andrew T. Campbell. Tracking Depression Dynamics in College Students Using Mobile Phone and Wearable Sensing. Proceedings of the ACM on Interactive, Mobile, Wearable and Ubiquitous Technologies, 2(1), 2018. doi: 10.1145/3191775.

22. Kennedy Opoku Asare, Isaac Moshe, Yannik Terhorst, Julio Vega, Simo Hosio, Harald Baumeister, Laura Pulkki-Råback, and Denzil Ferreira. Mood ratings and digital biomarkers from smartphone and wearable data differentiates and predicts depression status: A longitudinal data analysis. Pervasive and Mobile Computing, 83, 2022. ISSN 15741192. doi: 10.1016/j.pmcj.2022.101621.

23. Paola Pedrelli, Szymon Fedor, Asma Ghandeharioun, Esther Howe, Dawn F. Ionescu, Darian Bhathena, Lauren B. Fisher, Cristina Cusin, Maren Nyer, Albert Yeung, Lisa Sangermano, David Mischoulon, Johnathan E. Alpert, and Rosalind W. Picard. Monitoring Changes in Depression Severity Using Wearable and Mobile Sensors. Frontiers in Psychiatry, 11, 12 2020. ISSN 16640640. doi: 10.3389/fpsyt.2020.584711.

24. Robert A. Lewis, Asma Ghandeharioun, Szymon Fedor, Paola Pedrelli, Rosalind Picard, and David Mischoulon. Mixed Effects Random Forests for Personalised Predictions of Clinical Depression Severity. 1 2023.

25. Janik Goltermann, Daniel Emden, Elisabeth Johanna Leehr, Katharina Dohm, Ronny Redlich, Udo Dannlowski, Tim Hahn, and Nils Opel. Smartphone-based self-reports of depressive symptoms using the remote monitoring application in psychiatry (ReMAP): Interformat validation study. JMIR Mental Health, 8(1), 1 2021. ISSN 23687959. doi: 10.2196/24333.

26. Daniel Emden, Janik Goltermann, Udo Dannlowski, Tim Hahn, and Nils Opel. Technical feasibility and adherence of the Remote Monitoring Application in Psychiatry (ReMAP) for the assessment of affective symptoms. Journal of Affective Disorders, 294:652–660, 11 2021. ISSN 15732517. doi: 10.1016/j.jad.2021.07.030.

27. Martin Ester, Hans-Peter Kriegel, Jörg Sander, and Xiaowei Xu. A Density-Based Algorithm for Discovering Clusters in Large Spatial Databases with Noise. In Proceedings of the 2nd International Conference on Knowledge Discovery and Data Mining, 1996.

28. Habiballah Rahimi-Eichi, Garth Coombs, Jukka-Pekka Onnela, Justin T Baker, and Randy L Buckner. Measures of Behavior and Life Dynamics from Commonly Available GPS Data (DPLocate): Algorithm Development and Validation. 2022.

29. C Kühner, C Bürger, F Keller, and M Hautzinger. Reliabilität und Validität des revidierten Beck-Depressionsinventars (BDI-II) [Reliability and validity of the Revised Beck Depression Inventory (BDI-II). Results from German samples]. Der Nervenarzt, 78(6), 2007. ISSN 0028-2804.

30. Ramona Leenings, Nils Ralf Winter, Lucas Plagwitz, Vincent Holstein, Jan Ernsting, Kelvin Sarink, Lukas Fisch, Jakob Steenweg, Leon Kleine-Vennekate, Julian Gebker, Daniel Emden, Dominik Grotegerd, Nils Opel, Benjamin Risse, Xiaoyi Jiang, Udo Dannlowski, and Tim Hahn. PHOTONAI-A Python API for rapid machine learning model development. PLoS ONE, 16(July), 7 2021. ISSN 19326203. doi: 10.1371/journal.pone.0254062.

31. Colin R. Blyth. On simpson’s paradox and the sure-thing principle. Journal of the American Statistical Association, 67(338), 1972. ISSN 1537274X. doi: 10.1080/01621459.1972.10482387.

32. Kennedy Opoku Asare, Yannik Terhorst, Julio Vega, Ella Peltonen, Eemil Lagerspetz, and Denzil Ferreira. Predicting depression from smartphone behavioral markers using machine learning methods, hyperparameter optimization, and feature importance analysis: Exploratory study. JMIR mHealth and uHealth, 9(7), 7 2021. ISSN 22915222. doi: 10.2196/26540.

33. Shweta Ware, Chaoqun Yue, Reynaldo Morillo, Jin Lu, Chao Shang, Jinbo Bi, Jayesh Kamath, Alexander Russell, Athanasios Bamis, and Bing Wang. Predicting Depressive Symptoms Using Smartphone Data. 2019. doi: 10.1016/j.smhl.xxxx.xx.xxx.

34. Kelly M. Shaffer, Philip I. Chow, Jillian V. Glazer, Tri Le, Matthew J. Reilley, Mark J. Jameson, and Lee M. Ritterband. Feasibility of ecological momentary assessment to study depressive symptoms among cancer caregivers. Psycho-Oncology, 30(5), 2021. ISSN 10991611. doi: 10.1002/pon.5627.

35. Jonah Meyerhoff, Tony Liu, Konrad P. Kording, Lyle H. Ungar, Susan M. Kaiser, Chris J. Karr, and David C. Mohr. Evaluation of changes in depression, anxiety, and social anxiety using smartphone sensor features: Longitudinal cohort study. Journal of Medical Internet Research, 23(9), 2021. ISSN 14388871. doi: 10.2196/22844.

36. Mehmet Yücel Ag?argün, Hayrettin Kara, and Mustafa Solmaz. Subjective sleep quality and suicidality in patients with major depression. Journal of Psychiatric Research, 31(3), 1997. ISSN 00223956. doi: 10.1016/S0022-3956(96)00037-4.

37. Yasuaki Hayashino, Shin Yamazaki, Misa Takegami, Takeo Nakayama, Shigeru Sokejima, and Shunichi Fukuhara. Association between number of comorbid conditions, depression, and sleep quality using the Pittsburgh Sleep Quality Index: Results from a population-based survey. Sleep Medicine, 11(4), 2010. ISSN 13899457. doi: 10.1016/j.sleep.2009.05.021.

38. Yousef D. Alqurashi, Ali H. Al Qattan, Hassan E. Al Abbas, Majed A. Alghamdi, Abdullah A. Alhamad, Hashem A. Al-Dalooj, Talay Yar, Noor A. Al Khathlan, Abdullah S. Alqarni, and Ayad M. Salem. Association of sleep duration and quality with depression among university students and faculty. Acta Biomedica, 93(5), 2022. ISSN 25316745. doi: 10.23750/abm.v93i5.13002.

39. Avshalom Caspi and Terrie E. Moffitt. All for one and one for all: Mental disorders in one dimension. American Journal of Psychiatry, 175(9), 9 2018. ISSN 15357228. doi: 10.1176/appi.ajp.2018.17121383.

